# Influence of vitamin D supplementation on muscle strength and exercise capacity in South African schoolchildren: a randomised controlled trial (ViDiKids)

**DOI:** 10.1101/2024.03.26.24304912

**Authors:** Keren Middelkoop, Lisa K Micklesfield, Stephanie Hemmings, Neil Walker, Justine Stewart, David A Jolliffe, Amy E Mendham, Jonathan C Y Tang, Cyrus Cooper, Nicholas C Harvey, Robert J Wilkinson, Adrian R Martineau

**Author notes:** To whom correspondence should be addressed at the Blizard Institute, Barts and The London School of Medicine and Dentistry, Queen Mary University of London, 4 Newark St, London E1 2AT, UK.

## Abstract

**Objective:** To determine whether vitamin D supplementation influences grip strength, explosive leg power, cardiorespiratory fitness and risk of exercise-induced bronchoconstriciton (EIB) in South African schoolchildren.

**Methods:** Sub-study (n=450) in Cape Town schoolchildren aged 8-11 years, nested within a phase 3 randomised placebo-controlled trial (ViDiKids). The intervention was weekly oral doses of 10,000 IU vitamin D_3_ (n=228) or placebo (n=222) for 3 years. Outcome measures were serum 25-hydroxyvitamin D_3_ (25[OH]D_3_) concentrations, grip strength, standing long jump distance, peak oxygen uptake (VO_2peak_, determined using 20-metre multi-stage shuttle run tests) and the proportion of children with EIB, all measured at end-study.

**Results:** 64.7% of participants had serum 25(OH)D_3_ concentrations <75 nmol/L at baseline. At 3-year follow-up, children randomised to vitamin D vs. placebo had higher mean serum 25(OH)D_3_ concentrations (97.6 vs. 58.8 nmol/L respectively; adjusted mean difference [aMD] 39.9 nmol/L, 95% CI 36.1 to 43.6) and long jump distance (128.3 vs. 122.1 cm; aMD 3.6 cm, 95% CI 0.0 to 7.2). No end-study differences in grip strength, VO_2peak_, or spirometric lung volumes were seen, but administration of vitamin D vs. placebo was associated with a borderline-significant increased risk of EIB (14.5% vs. 8.6%; adjusted odds ratio 1.92, 95% CI 0.99 to 3.73).

**Conclusion:** A 3-year course of weekly oral supplementation with 10,000 IU vitamin D_3_ elevated serum 25(OH)D_3_ concentrations in South African schoolchildren and induced a small increase in long jump distance, but had no effect on grip strength or V_O2 peak_. Potential effects of vitamin D on risk of EIB require further research.

**KEY MESSAGES:** *What is already known on this topic?:* Observational studies have reported that vitamin D deficiency associates with reduced muscle strength and peak oxygen uptake and increased risk of exercise-induced bronchoconstriction (EIB) in children. Randomised controlled trials (RCT) of vitamin D supplementation to improve children’s muscle strength and cardiorespiratory fitness have yielded conflicting results.

*What this study adds:* This RCT, conducted in South African schoolchildren aged 8-11 years at baseline, found that a 3-year course of weekly oral supplementation with 10,000 IU vitamin D_3_ improved vitamin D status and resulted in a small (3.6 cm), borderline-significant increase in long jump distance, but had no effect on grip strength or peak oxygen uptake. Administration of vitamin D was associated with a borderline-significant increased risk of EIB.

*How this study might affect research, practice or policy:* Taken together with null results from another Phase 3 randomised controlled trial of vitamin D supplementation conducted in Mongolian children, our findings do not suggest that weekly oral vitamin D supplementation exerts clinically significant effects on muscle strength or peak oxygen uptake in schoolchildren in whom rickets has been excluded. Further research into potential effects of vitamin D supplementation on risk of EIB is needed.

## INTRODUCTION

Muscle strength and exercise tolerance positively correlate with physical and mental health during childhood, and associate with reduced risk of cardiometabolic disease in adulthood.^1–4^ Vitamin D is essential for the normal development and function of the cardiorespiratory system and skeletal muscle.^5–7^ Deficiency in this fat-soluble micronutrient – indicated by low circulating concentrations of 25-hydroxyvitamin D (25[OH]D) - is widespread among children in both higher- and lower-income countries.^8–11^ Observational studies have linked vitamin D deficiency in children and adolescents to reduced muscle strength and cardiorespiratory fitness,^12^ ^13^ and increased risk of exercise induced bronchoconstriction (EIB),^14^ which may limit exercise capacity. Numerous randomised controlled trials (RCTs) in adults have revealed positive effects of vitamin D supplementation on muscle strength and power, with meta-analyses reporting a modest benefit overall.^15^ However, analogous studies in children are less numerous and have yielded inconsistent outcomes. For example, a RCT of vitamin D supplementation conducted in Tunisia in vitamin D deficient male soccer players aged 8-15 years reported improvements in long jump distance, sprint and shuttle run outcomes,^16^ while a UK study in 12-14 year-old girls showed that vitamin D supplements enhanced movement efficiency, with indications of better jumping velocity and grip among participants randomised to intervention.^17^ However, RCTs in children and adolescents living in Denmark,^18^ ^19^ the United States,^20^ Israel,^21^ and Lebanon^22^ have not found significant effects of vitamin D supplementation on grip strength, leg press strength, or swimming performance. No such trials have yet been conducted in Africa; moreover, there is a lack of large, multicentre trials examining the impact of prolonged (greater than one year) vitamin D supplementation on muscle strength, cardiorespiratory fitness and risk of EIB in children living in any setting.

Recently, an opportunity to generate new data in this area arose in association with conduct of a phase 3 RCT of weekly vitamin D supplementation involving a total of 1,682 children attending primary schools in Cape Town, South Africa. The primary objective was to determine whether this intervention would decrease risk of incident tuberculosis infection, as indicated by conversion of an Interferon-Gamma Release Assay from negative at baseline to positive at 3-year follow-up; null findings for this outcome have been reported elsewhere.^23^ The current paper reports findings of a sub-study nested within the main trial, which investigated whether vitamin D supplementation influenced grip strength, standing long jump distance, peak oxygen uptake (VO_2peak_) or risk of exercise-induced bronchoconstriction in a subset of 450 participants aged 8-11 years at baseline.

## METHODS

### TRIAL DESIGN, SETTING, APPROVALS AND REGISTRATION

We conducted a multicentre phase 3 double-blind individually randomised placebo-controlled trial in 23 government schools in Cape Town, South Africa, as previously described.^23–25^ The primary outcome was acquisition of latent tuberculosis infection; the current manuscript reports effects of the intervention on pre-specified secondary outcomes relating to muscle strength and cardiorespiratory outcomes measured at 3-year follow-up in a subset of 450 participants who took part in a nested sub-study. The trial was sponsored by Queen Mary University of London and registered on the South African National Clinical Trials Register (DOH-27-0916-5527) and ClinicalTrials.gov (ref NCT02880982).

### PARTICIPANTS

Inclusion criteria for the main trial were enrolment in Grades 1-4 at a participating school; age 6 to 11 years at screening; and written informed assent / consent to participate in the main trial provided by children and their parent / legal guardian, respectively. Exclusion criteria for the main trial were a history of previous latent TB infection, active TB disease or any chronic illness other than asthma (including known or suspected HIV infection) prior to enrolment; use of any regular medication other than asthma medication; use of vitamin D supplements at a dose of more than 400 IU/day in the month before enrolment; plans to move away from study area within 3 years of enrolment; inability to swallow a placebo soft gel capsule with ease; and clinical evidence of rickets or a positive QFT-Plus assay result at screening. Additional inclusion criteria for the sub-study were enrolment in Grade 4 at a participating school; additional exclusion criteria for the sub-study were presence of an injury or disability limiting ability to run with maximum effort, and inability to perform spirometry at baseline.

### ENROLMENT AND BASELINE ASSESSMENTS

Parents or legal guardians were invited to provide written informed consent for their child to participate in the main trial during a home visit, unless their child was eligible for the sub-study, in which case they were invited to provide written informed consent for their child to participate in both the main trial and the sub-study until a total of 450 sub-study participants were randomised. If parents / legal guardians consented, they were asked to provide details of their child’s dietary intake of foods containing vitamin D and calcium in the previous month, which were captured on an electronic case report form as previously described.^24^ Their children were then invited to provide written assent to participate in the main trial +/- the sub-study (if eligible) at a school-based visit. If they agreed, a clinically trained member of the study team screened them for symptoms and signs of rickets. For all participants, a blood sample was taken for a QuantiFERON-TB Gold Plus (QFT-Plus) assay (Qiagen, Hilden, Germany) and separation and storage of serum for determination of 25(OH)D concentrations as described below. Participants were reviewed when baseline QFT-Plus results were available. Those with a positive QFT-Plus result were excluded from the trial and screened for active TB. Those with an indeterminate QFT-Plus result were excluded from the trial without screening for active TB. Those with a negative QFT-Plus result were deemed eligible to participate and underwent measurement of weight (using a digital floor scale, Charder Medical) and height (using a portable HM200P stadiometer, Charder Medical). For sub-study participants, grip strength was measured as described elsewhere^26^ using a portable dynamometer (Takei Digital Grip Strength Dynamometer, Model T.K.K.5401); the best of 2 readings for the dominant hand were recorded, except where injury precluded measurement, where strength of the non-dominant hand was measured. Standing long jump distance was measured as described elsewhere^27^ using a DiCUNO measuring tape, with the best of 2 readings recorded. Spirometry was performed according to ERS/ATS standards^28^ using a portable spirometer (SpiroUSB, Carefusion, San Diego, USA). A 20-meter multi-stage shuttle run test was then conducted using freely available recorded instruction (Shuttle run bleep test, www.bleeptests.com, accessed 26^th^ March 2024) until volitional exhaustion. Two lines were marked 20 metres apart and an audible ‘bleep’ signalled to participants the speed required to run between them. The number of completed laps was recorded and used to derive VO_2peak_ as described below. Spirometry was repeated 3-5 minutes after completion of the shuttle run test to detect EIB.

### RANDOMISATION AND BLINDING

Eligible and assenting children whose parents consented to their participation in the trial were individually randomised to receive a weekly capsule containing vitamin D_3_ or placebo for three years, with a one-to-one allocation ratio and randomisation stratified by school of attendance, as previously described.^23^ Treatment allocation was concealed from participants, care providers and all trial staff (including senior investigators and those assessing outcomes) until completion of the trial to maintain the double-blind.

### INTERVENTION

Study medication comprised a 3-year course of weekly soft gel capsules manufactured by the Tishcon Corporation (Westbury, NY, USA), containing either 0.25 mg (10,000 international units) cholecalciferol (vitamin D_3_) in olive oil (intervention arm) or olive oil without any vitamin D_3_ content (placebo arm). Active and placebo capsules had identical appearance and taste. Capsules were taken under direct observation of study staff during school termtime. During summer holidays (8 weeks), packs containing 8 doses of study medication were provided for administration by parents, together with a participant diary. Following shorter school holidays (≤4 weeks), and/or if participants missed one or more doses of study medication during term time, up to 4 ‘catch-up’ doses were administered at the first weekly visit attended following the missed dose(s). During the initial national lockdown for COVID-19 in South Africa (27^th^ March to 1^st^ May 2020), participants did not receive any study medication. During subsequent school closures due to COVID-19, two rounds of 8-week holiday packs were provided to participants, which were sufficient to cover their requirements until schools re-opened.

### OUTCOMES

The primary outcome for the main trial, reported elsewhere,^23^ was the QuantiFERON-TB Gold Plus result at the manufacturer-recommended 0.35 IU/mL threshold at the end of the study. The following secondary outcomes were assessed for sub-study participants at 3-year follow-up: maximal grip strength (kg), standing long jump distance (cm), VO_2peak_ (ml/kg/min, derived from shuttle run performance) and the proportion of participants with EIB, defined as a ≥10% decrease in forced expiratory volume in 1 second (FEV1) after completion vs. before the shuttle run test.^29^

### LABORATORY ASSESSMENTS

Biochemical analyses were performed at the Bioanalytical Facility, University of East Anglia (Norwich, UK) according to manufacturers’ instructions and under Good Clinical and Laboratory Practice conditions. Serum concentrations of 25(OH)D_3_ were measured using liquid chromatography tandem mass spectrometry (LC-MS/MS) as previously described.^10^ 25(OH)D_3_ was calibrated using standard reference material SRM972a from the National Institute of Science and Technology (NIST), and the assay showed linearity between 0 and 200 nmol/L. The inter/intra-assay coefficient of variation (CV) across the assay range was ≤9%, and the lower limit of quantification was 0.1 nmol/L. The assay showed <6% accuracy bias against NIST reference method on the vitamin D external quality assessment (DEQAS) scheme (http://www.deqas.org/; accessed on 18^th^ March 2024). QFT-Plus assays were performed by the Bio Analytical Research Corporation South Africa (Johannesburg, South Africa) according to the manufacturer’s instructions.

### SAMPLE SIZE

Sample size for the main trial was predicated on power to detect an effect of the intervention on the primary outcome (the proportion of children with a positive QFT-Plus assay result at 3-year follow-up), as previously described.^23^ Sample size for the sub-study was predicated on power to detect an effect of the intervention on bone mineral content, as previously described.^24^

### STATISTICAL ANALYSES

Statistical analyses were performed using Stata software (Version 17.0; StataCorp, College Station, Texas, United States) according to intention to treat. VO_2peak_ was calculated from the number of completed laps on shuttle run tests using a published formula.^30^ Effects of treatment on continuous outcomes measured at 3-year follow-up were analysed using multi-level mixed models with random effects of school and individual, and with the treatment effect at baseline constrained to be zero to reflect the randomisation, where applicable. Proportions of children with EIB at 3-year follow-up were analysed using a mixed-effects logistic regression model, with treatment allocation as the sole fixed effect and school attended as a random intercept and adjustment for presence vs absence of EIB at baseline. Pre-specified sub-group analyses were conducted to determine whether the effect of vitamin D supplementation was modified by sex (male vs. female), baseline deseasonalised 25(OH)D_3_ concentration, calculated using a sinusoidal model as previously described^31^ (<75 vs. ≥75 nmol/L) and estimated calcium intake (< vs. ≥ median value of 466 mg/day, calculated as previously described).^24^ These were performed by repeating efficacy analyses with the inclusion of an interaction term between allocation (to vitamin D vs. placebo) and each posited effect-modifier with presentation of the P-value associated with this interaction term. Given the number of potential effect modifiers and secondary outcome measures these analyses are considered exploratory. Interim safety assessments, where Independent Data Monitoring Committee (IDMC) members reviewed accumulating serious adverse event data, were performed at 6-monthly intervals. At each review the IDMC recommended continuation of the trial. No interim efficacy analysis was performed.

### PATIENT AND PUBLIC INVOLVEMENT

The ViDiKids trial was designed in consultation with the local Community Advisory Board (CAB) – a committee of local community stakeholders. Plans for enrolment and implementation of the study protocol were discussed with the school principles, staff and parent bodies prior to study initiation. Collaborative relationships with participating schools meant that the staff assisted in addressing study challenges, especially those that arose as a result of the COVID-19 pandemic. Dissemination of study results to the CAB, the school staff and parent bodies by a team including a staff member fluent in the local language is on-going.

## RESULTS

### PARTICIPANTS

2852 children were screened for eligibility to participate in the main trial from March 2017 to March 2019, of whom 2271 underwent QFT testing: 1682 (74.1%) QFT-negative children were randomly assigned to receive vitamin D_3_ (829 participants) or placebo (853 participants) as previously described.^23^ 450/1682 (26.8%) participants in the main trial also participated in the sub-study, of whom 228 vs. 222 participants were allocated to the vitamin D vs. placebo arms, respectively (Fig. 1). Table 1 shows baseline characteristics of participants contributing data to analyses of cardiorespiratory and strength outcomes: overall, mean age was 10.1 years, 52.0% were female and mean deseasonalised baseline serum 25(OH)D concentration was 70.0 nmol/L (s.d. 13.5 nmol/L). These and other baseline characteristics were well-balanced between sub-study participants randomised to vitamin D vs. placebo. Of 450 randomised sub-study participants, 391 (86.9%) attended the sub-study follow-up visit at 3 years post-randomisation and contributed data to analyses presented here. Mean serum 25(OH)D_3_ concentration at 3-year follow-up was higher in participants randomised to vitamin D vs. placebo (97.6 vs. 58.8 nmol/L, respectively: mean difference 39.9 nmol/L, 95% CI, 36.1 to 43.6).

**Figure 1.**
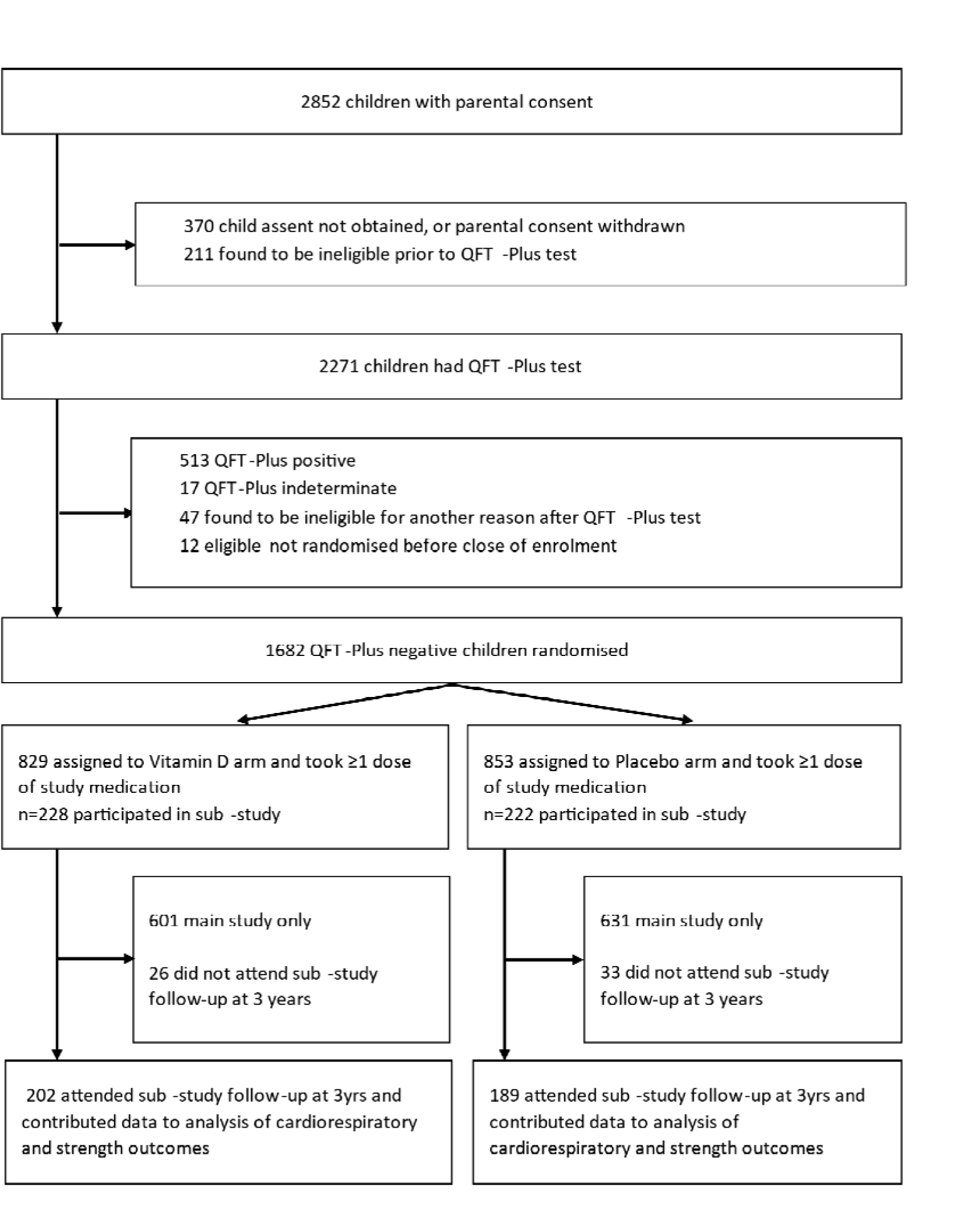
Participant Flow Diagram

**Table 1:** Participants’ baseline characteristics, overall and by allocation.

|  |  | Overall<br>(n=450) | Vitamin D arm<br>(n=228) | Placebo arm<br>(n=222) |
| --- | --- | --- | --- | --- |
| Mean age, years (s.d.) [range] |  | 10.1 (0.7) [8.2 to 11.9] | 10.2 (0.7) [8.2 to 11.9] | 10.0 (0.6) [8.5 to 11.9] |
| Sex, n (%) | Female | 234 (52.0) | 116 (50.9) | 118 (53.2) |
|  | Male | 216 (48.0) | 112 (49.1) | 104 (46.8) |
| Ethnic origin <sup>(1)</sup> | Xhosa, n (%) | 424 (96.4) | 214 (96.0) | 210 (96.8) |
|  | Other, n (%) | 16 (3.6) | 9 (4.0) | 7 (3.2) |
| Type of residence | Brick, n (%) | 230 (51.1) | 121 (53.1) | 109 (49.1) |
|  | Informal, n(%) | 220 (48.9) | 107 (46.9) | 113 (50.9) |
| Parental education <sup>(2)</sup> | None, n(%) | - | - | - |
|  | Primary school, n (%) | 24 (5.3) | 16 (7.0) | 8 (3.6) |
|  | Secondary school or higher, n (%) | 426 (94.7) | 212 (93.0) | 214 (96.4) |
| Mean monthly household income, 1,000 ZAR (s.d.) |  | 1.6 (2.3) | 1.5 (2.7) | 1.6 (1.9) |
| Household environmental tobacco smoke, n (%) |  | 47 (10.4) | 28 (12.3) | 19 (8.6) |
| Mean maximal grip strength, kg (s.d.) <sup>(3)</sup> |  | 13.9 (3.4) | 14.2 (3.6) | 13.7 (3.1) |
| Mean standing long jump distance, cm (s.d.) <sup>(3)</sup> |  | 120.0 (18.4) | 121.7 (18.5) | 118.2 (18.1) |
| Mean VO<sub>2peak</sub>, ml/kg/min (s.d.) <sup>(3)</sup> |  | 40.5 (3.2) | 40.5 (3.0) | 40.5 (3.3) |
| Exercise-induced bronchospasm, proportion (%) <sup>(3)</sup> |  | 79/448 (17.6) | 38/228 (16.7) | 41/220 (18.6) |
| Mean serum 25(OH)D<sub>3</sub>, nmol/L (s.d.) [range] <sup>(4)</sup> |  | 70.0 (13.5) [24.9 to 112.5] | 70.4 (12.1) [43.2 to 101.3] | 69.6 (14.9) [24.9 to 112.5] |
| Serum 25(OH)D<sub>3</sub> categories, n (%) <sup>(4)</sup> | <25 nmol/L | 1 (0.3) | 0 (0.0) | 1 (0.6) |
|  | 25-49.9 nmol/L | 20 (5.5) | 7 (3.7) | 13 (7.4) |
|  | 50-74.9 nmol/L | 214 (59.0) | 114 (61.0) | 100 (56.8) |
|  | ≥75 nmol/L | 128 (35.3) | 66 (35.3) | 62 (35.2) |

**Footnotes:** (1) Ethnicity data missing in 5 participants in both the vitamin D and placebo arms. (2) Highest level of education of at least one parent. (3) Data missing in 2 participants in the placebo arm. (4) Serum 25(OH)D<sub>3</sub> concentrations deseasonalised; data missing in 41 participants in the vitamin D arm and 46 participants in the placebo arm.
**Abbreviations:** ZAR, South African Rand. VO<sub>2peak</sub>, peak oxygen consumption. FEV1, forced expiratory volume in 1 second. FVC, forced vital capacity. 25(OH)D<sub>3</sub>, 25-hydroxyvitamin D<sub>3</sub>. S.d., standard deviation.

### OUTCOMES

Allocation to vitamin D vs. placebo did not influence grip strength, either overall or within sub-groups defined by male vs. female sex, baseline 25(OH)D concentration or calcium intake (Table 2). By contrast, participants randomised to vitamin D vs. placebo had a slightly higher mean long jump distance at 3-year follow-up (128.2 vs. 122.1 cm, respectively; adjusted mean difference 3.6 cm, 95% CI 0.0 to 7.2 cm; P=0.0502). Sub-group analyses revealed no evidence of effect-modification by sex, baseline 25(OH)D concentration or calcium intake for this outcome (Table 2). No effect of allocation to vitamin D vs. placebo was seen for mean VO_2peak_, either overall or by sub-group (Tables 3). However, a borderline statistically significant difference in the proportion of participants with EIB was seen between participants allocated to vitamin D vs. placebo (14.5% vs. 8.6% respectively, aOR 1.92, 95% CI 0.99 to 3.73; P=0.053; Table 4).

**Table 2.** Maximal grip strength and standing long jump distance at 3-year follow-up by allocation: overall and by sub-groups.

|  |  | Vitamin D: mean value, kg (s.d.) [n] | Placebo: mean value, kg (s.d.) [n] | Adjusted mean difference (95% CI) <sup>(1)</sup> | P | P for interaction |
| --- | --- | --- | --- | --- | --- | --- |
| **Grip strength** |  |  |  |  |  |  |
| Overall |  | 20.9 (5.1) [201] | 20.6 (5.2) [188] | -0.19 (-0.94 to 0.55) | 0.61 | -- |
| By sex | Male | 22.0 (5.5) [96] | 21.3 (5.9) [89] | 0.04 (-1.24 to 1.32) | 0.95 | 0.57 |
|  | Female | 19.8 (4.4) [105] | 20.0 (4.4) [99] | -0.38 (-1.22 to 0.46) | 0.37 |  |
| By baseline 25(OH)D concentration <sup>(2)</sup> | <75 nmol/L | 20.6 (4.7) [103] | 21.1 (5.3) [96] | -0.51 (-1.58 to 0.56) | 0.35 | 0.14 |
|  | ≥75 nmol/L | 21.7 (6.2) [60] | 20.5 (5.3) [54] | 0.74 (-0.64 to 2.12) | 0.30 |  |
| By calcium intake | <median <sup>(3)</sup> | 20.9 (4.6) [93] | 20.9 (5.5) [97] | -0.25 (-1.43 to 0.94) | 0.68 | 0.65 |
|  | ≥median <sup>(3)</sup> | 21.0 (5.6) [102] | 20.5 (4.8) [88] | -0.12 (-1.02 to 0.78) | 0.79 |  |
|  |  | Vitamin D: mean value, cm (s.d.) [n] | Placebo: mean value, cm (s.d.) [n] | Adjusted mean difference (95% CI) <sup>(1)</sup> | P | P for interaction |
| **Long jump distance** |  |  |  |  |  |  |
| Overall |  | 128.2 (22.1) [201] | 122.1 (21.0) [188] | 3.58 (-0.00 to 7.16) | 0.0502 | -- |
| By sex | Male | 140.5 (20.3) [96] | 132.7 (20.2) [89] | 5.89 (0.80 to 10.99) | 0.02 | 0.32 |
|  | Female | 117.0 (17.2) [105] | 112.6 (17.0) [99] | 2.58 (-1.47 to 6.62) | 0.21 |  |
| By baseline 25(OH)D concentration <sup>(2)</sup> | <75 nmol/L | 130.0 (24.4) [93] | 124.7 (20.7) [97] | 1.14 (-3.76 to 6.05) | 0.65 | 0.62 |
|  | ≥75 nmol/L | 126.7 (19.3) [102] | 119.9 (21.2) [88] | 3.66 (-2.54 to 9.86) | 0.25 |  |
| By calcium intake | <median <sup>(3)</sup> | 129.97 (24.39) [93] | 124.70 (20.72) [97] | 3.33 (-1.67 to 8.33) | 0.19 | 0.98 |
|  | ≥median <sup>(3)</sup> | 126.73 (19.35) [102] | 119.86 (21.21) [88] | 3.86 (-1.32 to 9.03) | 0.14 |  |

**Footnotes:** (1) adjusted for baseline value and school of attendance. (2) Serum 25(OH)D<sub>3</sub> concentrations deseasonalised. (3) median value 466 mg/day.
**Abbreviations:** 25(OH)D<sub>3</sub>, 25-hydroxyvitamin D<sub>3</sub>. CI, confidence interval. S.d., standard deviation. N, number.

**Table 3.** VO_2peak_ at 3-year follow-up by allocation: overall and by sub-groups.

|  |  | Vitamin D arm:<br>mean value,<br>ml/kg/min (s.d.) [n] | Placebo arm:<br>mean value,<br>ml/kg/min (s.d.)<br>[n] | Adjusted mean<br>difference (95% CI) <sup>(1)</sup> | P | P for<br>interaction |
| --- | --- | --- | --- | --- | --- | --- |
| Overall |  | 37.2 (4.1) [200] | 37.1 (4.1) [187] | 0.24 (-0.15 to 0.63) | 0.23 |  |
| By sex | Male | 39.0 (3.2) [95] | 39.2 (3.4) [89] | -0.02 (-0.56 to 0.53) | 0.95 | 0.22 |
|  | Female | 35.5 (4.1) [105] | 35.2 (3.8) [98] | 0.47 (-0.06 to 1.00) | 0.08 |  |
| By baseline 25(OH)D<br>concentration <sup>(2)</sup> | <25 nmol/L | 36.6 (4.5) [103] | 36.7 (3.9) [95] | 0.50 (-0.06 to 1.06) | 0.08 | 0.28 |
|  | ≥25 nmol/L | 37.7 (3.9) [59] | 37.5 (4.2) [54] | -0.01 (-0.75 to 0.74) | 0.98 |  |
| By calcium intake | <median <sup>(3)</sup> | 37.5 (3.7) [92] | 37.4 (4.3) [97] | 0.19 (-0.32 to 0.70) | 0.47 | 0.87 |
|  | ≥median <sup>(3)</sup> | 36.8 (4.5) [102] | 36.6 (4.0) [87] | 0.25 (-0.36 to 0.87) | 0.42 |  |

**Footnotes:** (1) adjusted for baseline value and school of attendance. (2) Serum 25(OH)D<sub>3</sub> concentrations deseasonalised. (3) median value 466 mg/day.
**Abbreviations:** 25(OH)D<sub>3</sub>, 25-hydroxyvitamin D<sub>3</sub>. Cm, centimeter. CI, confidence interval. S.d., standard deviation. N, number.

**Table 4.** Proportion of participants with exercise-induced bronchocontriction at 3-year follow-up by allocation: overall and by sub-group.

|  |  | Vitamin D:<br>proportion (%) | Placebo:<br>proportion (%) | Adjusted OR (95% CI)<br>(<sup>1</sup>) | P | P for<br>interaction |
| --- | --- | --- | --- | --- | --- | --- |
| Overall |  | 29/200 (14.50) | 16/187 (8.56) | 1.92 (0.99 to 3.73) | 0.053 |  |
| By sex | Male | 13/95 (13.68) | 5/89 (5.62) | 2.86 (0.95 to 8.61) | 0.06 | 0.32 |
|  | Female | 16/105 (15.24) | 11/98 (11.22) | 1.48 (0.65 to 3.41) | 0.35 |  |
| By baseline 25(OH)D<br>concentration (<sup>2</sup>) | <75 nmol/L | 14/103 (13.59) | 8/95 (8.42) | 1.82 (0.70 to 4.72) | 0.22 | 0.39 |
|  | ≥75 nmol/L | 7/59 (11.86) | 2/54 (3.70) | 3.49 (0.69 to 17.68) | 0.13 |  |
| By calcium intake | <median(<sup>3</sup>) | 13/92 (14.13) | 4/97 (4.12) | 3.86 (1.17 to 12.70) | 0.03 | 0.08 |
|  | ≥median(<sup>3</sup>) | 14/102 (13.73) | 12/87 (13.79) | 1.04 (0.44 to 2.46) | 0.93 |  |

**Footnotes:** (1) adjusted for school of attendance and presence vs. absence of exercise-induced bronchoconstriction at baseline. (2) deseasonalised values. (3) median value 466 mg/day.
**Abbreviations:** 25(OH)D, 25-hydroxyvitamin D. N, number. S.d., standard deviation. OR, odds ratio. CI, confidence interval.

## DISCUSSION

We present findings of the first RCT investigating effects of vitamin D supplementation on muscle strength and cardiorespiratory fitness to be conducted in Africa – and the first RCT conducted anywhere to test whether vitamin D supplementation influences risk of exercise-induced bronchoconstriction. Weekly oral administration of 10,000 IU vitamin D_3_ was effective in elevating circulating 25(OH)D concentrations: this was associated with a small, borderline-statistically significant increase in long jump distance, and a borderline-statistically significant increase in the proportion of participants with EIB at 3-year follow-up. No statistically significant inter-group differences were seen for other outcomes investigated.

Our null findings for grip strength and VO_2peak_ are in keeping with those from a similarly designed Phase 3 RCT conducted in Mongolia, where effects of a 3-year course of 14,000 IU vitamin D/week were investigated.^32–35^ Our finding of a small borderline-significant positive effect of vitamin D supplementation on long jump distance supports the concept that vitamin D has a physiological role in supporting muscle strength.^5^ However, the fact that this was not associated with an effect on grip strength, plus the observation that no such effect was seen in Mongolian children (who had a significantly higher baseline prevalence of vitamin D deficiency than participants in the current study)^10^ ^11^ suggests that this result may have arisen as a result of type 1 error arising as a result of investigating multiple secondary outcomes. The other borderline-significant finding was in the opposite direction to that anticipated: vitamin D supplementation associated with an increased risk of EIB at 3-year follow-up, in contrast to expectations that the intervention would reduce this risk via its immunoregulatory actions.^36^ The lack of any associated deleterious effect of vitamin D on VO_2peak_ in the current trial raises the possibility that this finding may also be attributable to type I error.

Our study has several strengths. Weekly vitamin D supplementation allowed direct observation of capsule administration during term-time, and this was reflected in the significant inter-arm difference in serum 25(OH)D concentrations observed at 3-year follow-up. 25(OH)D concentrations were measured using the gold standard methodology (LC-MS/MS) in a laboratory that participated in at rigorous external quality assessment scheme (DEQAS). Low rates of loss to follow-up maximised our power to detect effects of the intervention, and we assessed a comprehensive range of outcomes relating to muscle strength, cardiorespiratory fitness and respiratory function.

Our study also has some limitations. Fewer than 6% of children had baseline 25(OH)D concentrations <50 nmol/L, limiting generalisability of our findings to settings where vitamin D deficiency is more common. Additionally, we draw attention to that fact that our findings relate specifically to the effects of weekly vitamin D supplementation. Daily administration of vitamin D has been reported to be more beneficial for prevention of acute respiratory infections than weekly administration,^37^ and the possibility that this may also be the case for musculoskeletal and cardiorespiratory outcomes cannot be excluded. We also acknowledge that the parameters investigated in the current study were secondary outcomes (albeit pre-specified ones), and that multiple tests for statistical significance were performed; as discussed above, we cannot therefore rule out type I error as an explanation for statistically significant findings where these arose. In the absence of validated equations to estimate VO_2peak_ from shuttle run performance in South African children, we utilised equations developed for East Asian children, in order to maximise comparability of our findings with those of a similarly desgined RCT conducted in Mongolia.^32^ While we acknowledge that this may have introduced a degree of imprecision, randomisation will very likely have distributed this equally between intervention vs. control arms, minimising any risk of bias.

In conclusion, we report that a weekly oral dose of 14,000 IU vitamin D_3_, administered to South African children for 3 years, induced a small, borderline statistically significant increase in long jump distance, but did not influence grip strength or VO_2peak_. Administration of vitamin D was also associated with a borderline-significant increased risk of EIB; further research is needed to clarify potential effects of vitamin D supplementation on this outcome.

## Acknowledgements

We thank all the children who participated in the trial, and their parents / guardians; members of the Independent Data Monitoring Committee (Prof Guy Thwaites, Oxford University Clinical Research Unit, Ho Chi Minh City, Vietnam [Chair]; Prof John Pettifor, University of the Witwatersrand, Johannesburg, South Africa; and Prof Sarah Walker, MRC Clinical Trials Unit, London, UK); and members of the Trial Steering Committee (Prof Beate Kampmann, London School of Hygiene and Tropical Medicine, London, UK [Chair]; Prof Ashraf Coovadia, University of the Witwatersrand, Johannesburg, South Africa; Dr Karen Jennings, City Health, Cape Town, South Africa; Dr Robin Dyers, Western Cape Government, Cape Town, South Africa; and Dr Guy de Bruyn, Sanofi Pasteur, Swiftwater PA USA).

## Contributors

ARM conceived the study. KM, LKM, SH, CC, NCH, RJW and ARM contributed to study design and protocol development. KM led on trial implementation, with support from LKM, JS, DAJ, AEM, and ARM. JCYT performed and supervised the conduct of biochemical assays. NW and ARM drafted the statistical analysis plan. DAJ, KM, JS and NW managed data. NW, KM and DAJ accessed, verified and analysed the data underlying the study. NW and DAJ analysed data. ARM and DAJ wrote the first draft of the trial report. All authors made substantive comments thereon and approved the final version for submission.

## Patient consent for publication

Not applicable.

## Ethics approval

The trial was approved by the University of Cape Town Faculty of Health Sciences Human Research Ethics Committee (Ref: 796/2015) and the London School of Hygiene and Tropical Medicine Observational/Interventions Research Ethics Committee (Ref: 7450-2). Participants and their parents/guardians provided written informed assent and consent, respectively, to take part in the trial before any study procedures were conducted.

## Data availability statement

Anonymised data are available from corresponding authors upon reasonable request, subject to terms of IRB and regulatory approval.

